# Initial Simulation of SARS-CoV2 Spread and Intervention Effects in the Continental US

**DOI:** 10.1101/2020.03.21.20040303

**Authors:** Sen Pei, Jeffrey Shaman

## Abstract

Here we use a metapopulation model applied at county resolution to simulate the spread and growth of COVID-19 incidence in the continental United States. We calibrate the model against county-level incidence data collected between February 21, 2020 and March 13, 2020, and estimate epidemiological parameters including the fraction of undocumented infections and their contagiousness^1^. Using the calibrated model, we project the outbreak in the continental US for 180 days after March 13, 2020, and evaluate the effects of social distancing and travel restrictions on the outbreak.

## Model

We use a metapopulation SEIR model to simulate the transmission of COVID-19 among 3,108 US counties. In this model, we consider two types of movement: daily work commuting and random movement. Information on county-to-county work commuting is publicly available from the US Census Bureau^2^. We further assume the number of random visitors between two counties is proportional to the average number of commuters between them. As population present in each county is different at daytime and nighttime, we model the transmission dynamics of COVID-19 separately. We recognize that in the days leading up to March 13, some control measures were implemented in some areas of the country (e.g. school closure, closing of restaurants and bars, etc.); while these measures likely reduced inter-county movement to some degree, these effects were not likely large during this time period.

We formulate the transmission as a discrete Markov process during both day and night times. The daytime transmission lasts for *dt*_1_ day and the nighttime transmission *dt*_2_ day (*dt*_1_ + *dt*_2_ = 1). Here, we assume daytime transmission lasts for 8 hours and nighttime transmission lasts for 16 hours, i.e., *dt*_1_ = 1/3 and *dt*_2_ = 2/3. The transmission dynamics are depicted by the following equations.

Daytime transmission:

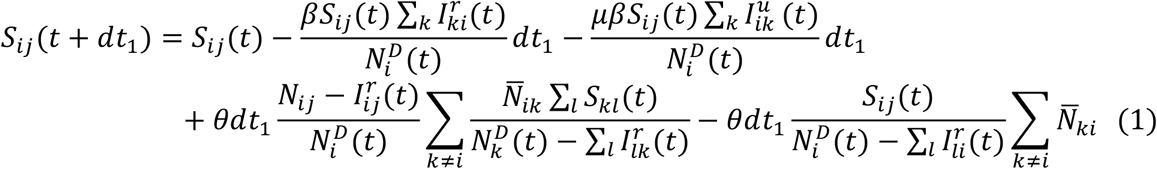

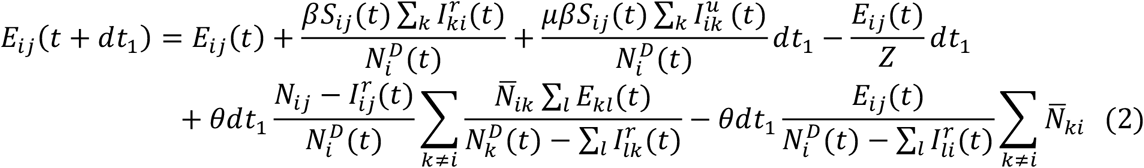

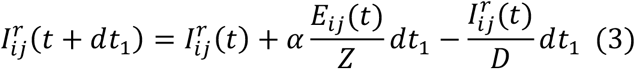

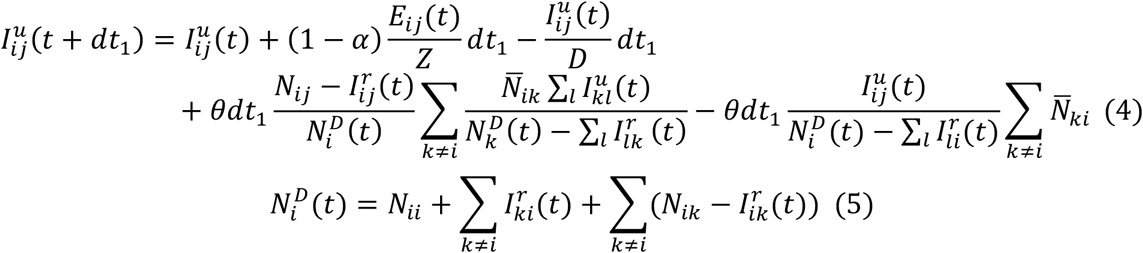

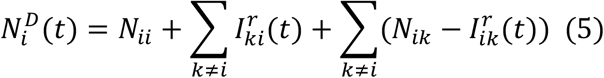

Nighttime transmission:

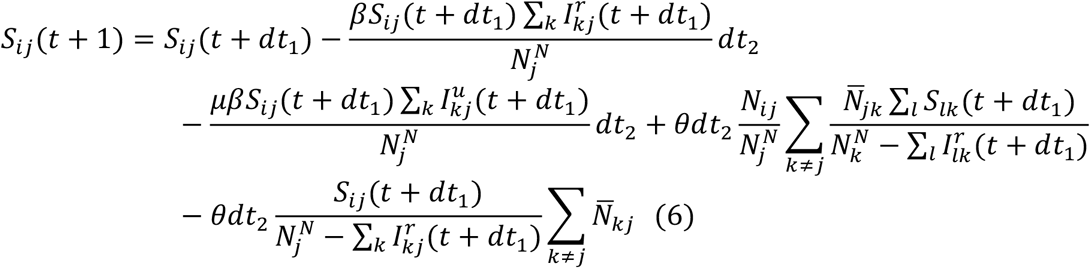

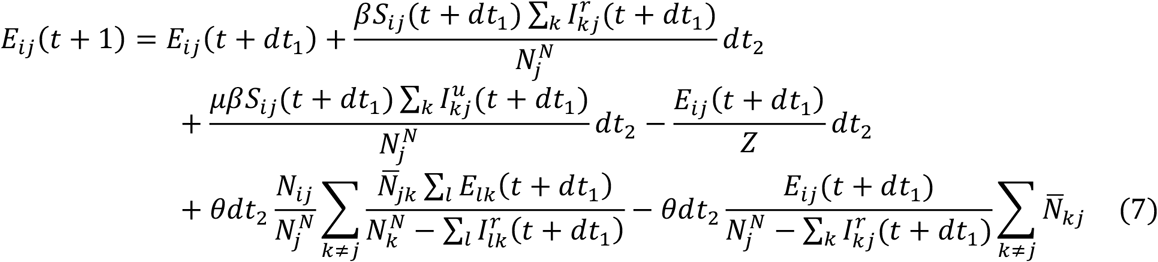

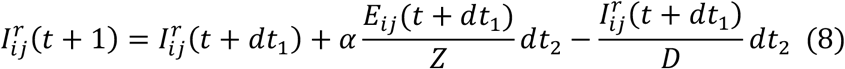

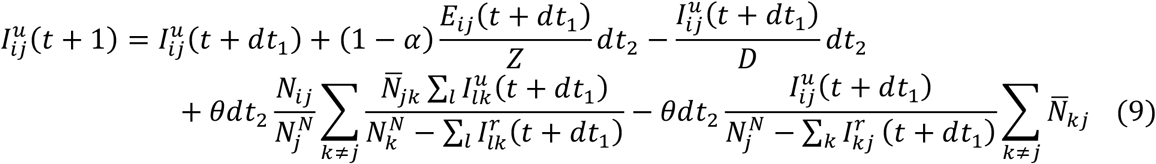

Here, 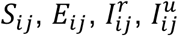 and *N*_*ij*_ are the susceptible, exposed, reported infected, unreported infected and total population in the subpopulation commuting from county *j* to county *i* (*i* ← *j*); *β* is the transmission rate of reported infections; *μ* is the relative transmissibility of unreported infections; *Z* is the average latency period (from infection to contagiousness); *D* is the average duration of contagiousness; *α* is the fraction of documented infections; *θ* is a multiplicative factor adjusting random movement; 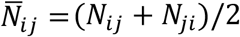 is the average number of commuters between counties *i* and *j*; and 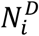 and 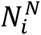 are the daytime and nighttime populations of county *i*. We integrate Eqs. 1-10 using a Poisson process to represent the stochasticity of the transmission process. A similar model has been used to generate forecasts for influenza in the United States^3^.

To account for reporting delay, we mapped simulated documented infections to confirmed cases using a separate observational delay model. In this delay model, we account for the time interval between a person transitioning from latent to contagious (i.e. 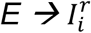) and observational confirmation of that individual infection. To estimate this delay period, *T*_*d*_, we examined line-list data from early-confirmed cases in China^4^. Prior to January 23, 2020, the time-to-event distribution of the interval (in days) from symptom onset to confirmation is well fit by a Gamma distribution^1^ (*a* = 1.85, *b* = 3.57, *LL* = −252.24). Consequently, we adopted a Gamma distribution to model *T*_*d*_, but tested longer mean periods (*ab*) as symptom onset often lags the onset of contagiousness.

### Parameter inference

We calibrated the transmission model against county-level incidence data reported from February 21, 2020 through March 13, 2020^5^. Specifically, we estimated model parameters using an iterated filtering (IF) framework^6,7^. The metapopulation model is high dimensional with 59,998 subpopulations. We therefore applied an efficient data assimilation algorithm – the Ensemble Adjustment Kalman Filter (EAKF)^8^, which is applicable to high dimensional models, in multiple iterations to infer parameters *β, μ, Z, D, α* and *θ*. This iterated filtering (IF)-EAKF framework has been used to infer parameters in a large-scale agent-based model for antimicrobial-resistant pathogens^9^, as well as a metapopulation model depicting the spread of SARS-CoV-2 in China^1^. Details of its implementation can be found in Ref. 1.

The prior ranges of model parameters were set as: *β* ∈ [0.3, 1.5], *μ* ∈ [0.2, 1.0], *Z* ∈ [2, 5], *D* ∈ [2, 5], *α* ∈ [0.02, 1.0], and *θ* ∈ [0.01,0.3]. In the inference, we fixed the shape parameter of the Gamma distribution for *T*_*d*_ as *a* = 1.85, and vary the mean value of the distribution. We tested a range of mean *T*_*d*_ values from 6 days to 10 days.

To initialize the model, we seeded exposed individuals (*E*) and unreported infections (*I*^*u*^) in counties with at least one confirmed case. Unlike the situation in China, where the outbreak originated from a single city, there was importation to multiple locations in the US that could have initiated community transmission. To reflect this potential ongoing community transmission before the reporting of the first local infection, for each county with confirmed cases, we randomly drew *E* and *I*^*r*^ from uniform distributions [0, 12*C*] and [0, 10*C*] 8 days prior to the reporting date (*T*_0_) of the first case. Here *C* is the total number of reported cases between day *T*_0_ and *T*_0_ + 4.

The rationale for this seeding strategy is as follows. If an average reporting delay of 8 days is assumed, we can estimate *I*^*r*^ on day *T*_0_ − 8 by 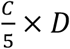, where 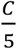 is the average number of daily cases during the first five days with reporting (*T*_0_ to *T*_0_ + 4). If we use the upper bound of the prior for D (i.e., 5 days), *I*^*r*^ is estimated as *C*, which is also an upper bound. Using parameters obtained from China^1^, we assume the mean *I*^*u*^ on day *T*_0_ − 8 is 5*C*, implying a reporting rate of 1/6=16.7%. Drawing *I*^*u*^ from [0, 10*C*] leads to a broader prior range of the reporting rate. As both *I*^*r*^ and *I*^*u*^ were evolved from the exposed population *E*, we draw *E* from the range [0, 12*C*]. This crude calculation gives us an estimate of seeding in US counties. During inference, this seeding can be adjusted up or down by the filter, and best-fitting models produce simulations that capture observed outcomes.

## Results

The inferred parameters for the best-fit model and the 95% CIs are reported in Table 1. The mean reporting delay for the best-fit model is *T*_*d*_ = 6 days. We display the fitting of this model to national daily case data and cases for 5 other counties (King County WA, Westchester County NY, Santa Clara County CA, Middlesex County MA and Nassau County NY) in Fig. 1.

**Table 1.** Best-fit model posterior estimates of key epidemiological parameters.

| Parameter | Median (95% CIs) |
| --- | --- |
| Transmission rate ($\beta$, days<sup>-1</sup>) | 0.95 (0.84, 1.06) |
| Relative transmission rate ($\mu$) | 0.64 (0.56, 0.70) |
| Latency period ($Z$, days) | 3.59 (3.28, 3.99) |
| Infectious period ($D$, days) | 3.56 (3.21, 3.83) |
| Reporting rate ($\alpha$) | 0.080 (0.069, 0.093) |
| Basic reproductive number ($R_e$) | 2.27 (1.87, 2.55) |
| Mobility factor ($\theta$) | 0.15 (0.12, 0.17) |

**Fig. 1.**
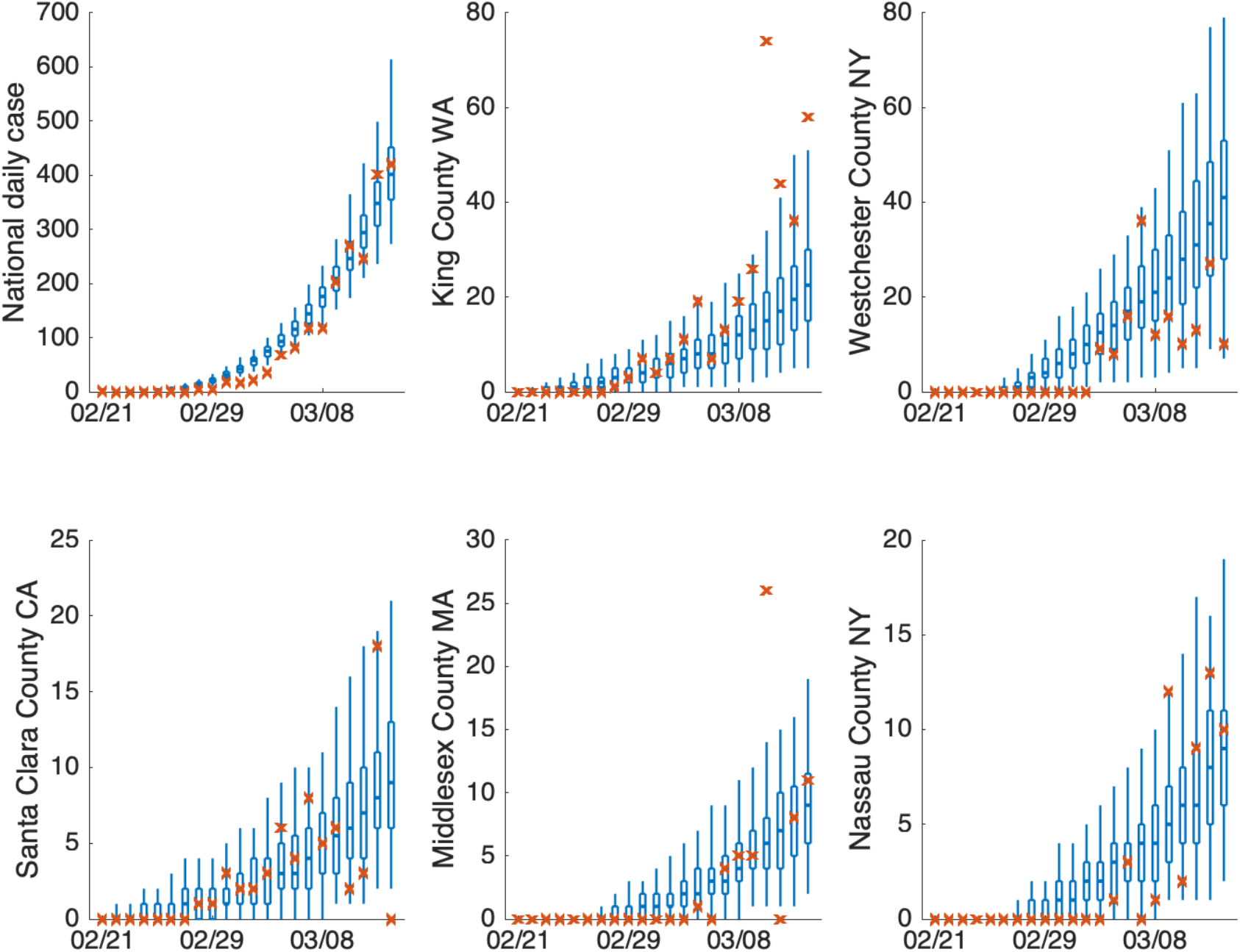
Model fitting to daily case from February 21 2020 to March 13 2020 in US, King County WA, Westchester County NY, Santa Clara County CA, Middlesex County MA and Nassau County NY.

To evaluate the impact of control measures, we examined how daily confirmed cases within 6 months of February 21 2020 were modulated by percent reductions of the contact rate *β* (effected by isolation, quarantine, telecommuting, school closure, etc.) and travel restrictions (effected by reducing commuting and travel among counties). The reductions of contact rates disrupt normal mixing within metapopulation locations, whereas the travel restrictions impact mixing between locations. In particular, we ran model simulations with no intervention, a 25% reduction of contact rate, a 50% reduction of contact rate and a 95% reduction of cross-county mobility. The epidemic curve is more effectively flattened by reduction of the contact rate, as shown in Fig. 2. In addition, we provide movies of each scenario (Movies S1-S4^10^). Each shows a single stochastic simulation with best-fitting model parameters (excepting *β*, which is adjusted in 2 of the scenarios). The projection outputs are available for download from GitHub^10^.

**Fig. 2.**
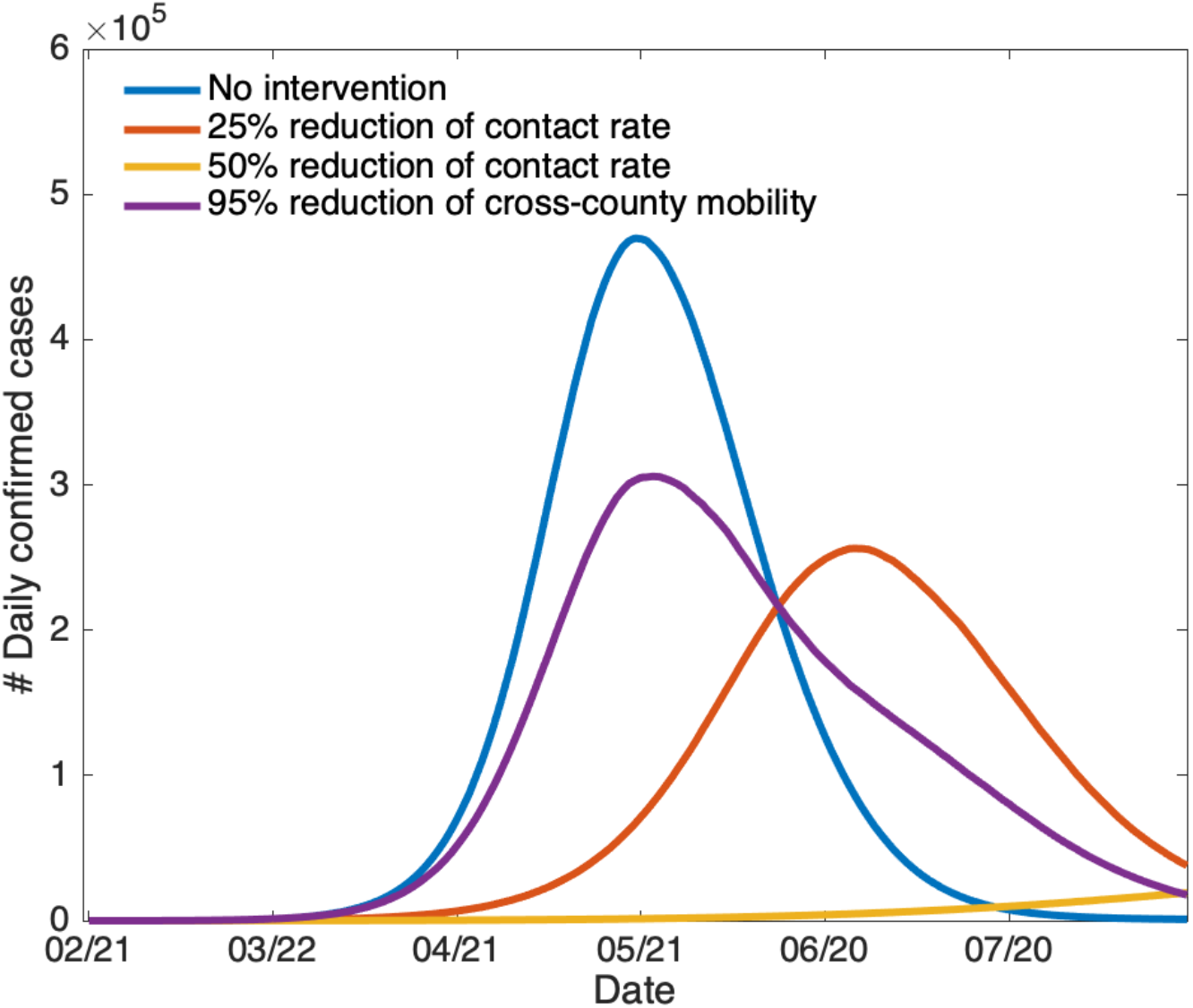
National daily confirmed cases with no intervention, a 25% reduction of contact rate, a 50% reduction of contact rate and a 95% reduction of cross-county mobility.

Disruption of mixing within locations, effected through a reduction of β, more substantially slows the spread and increase of confirmed cases than reductions of commuting and travel between locations. High reductions of commuting and travel between county locations (i.e. ≥95%) are needed to reduce the spread and increase of infections.

## Data Availability

Data are all publicly posted on Github and via the NY Times.

http://doi.org/10.5281/zenodo.3722660

https://www.nytimes.com/interactive/2020/us/coronavirus-us-cases.html

## References

1. Li R, Pei S, Chen B, Song Y, Zhang T, Yang W, Shaman J (2020) Substantial undocumented infection facilitates the rapid dissemination of novel coronavirus (SARS-CoV2). medRxiv, https://doi.org/10.1101/2020.02.14.20023127

2. https://www.census.gov/topics/employment/commuting.html

3. Pei S, Kandula S, Yang W, Shaman J. Forecasting the spatial transmission of influenza in the United States. PNAS 115(11): 2752–7 (2018).

4. M. Kramer, D. Pigott, B. Xu, S. Hill, B. Gutierrez, O. Pybus, Epidemiological data from the nCoV-2019 Outbreak: Early Descriptions from Publicly Available Data. Available: http://virological.org/t/epidemiological-data-from-the-ncov-2019-outbreak-early-descriptions-from-publicly-available-data/337 Accessed Feb 24, 2020.

5. https://www.nytimes.com/interactive/2020/us/coronavirus-us-cases.html. Accessed March 13 2020.

6. Ionides EL, Bretó C, King AA. Inference for nonlinear dynamical systems. PNAS 103(49):18438–43 (2006).

7. King AA, Ionides EL, Pascual M, Bouma MJ. Inapparent infections and cholera dynamics. Nature 454(7206):877–80 (2008).

8. Anderson JL. An ensemble adjustment Kalman filter for data assimilation. Monthly Weather Review 129(12):2884–903 (2001).

9. Pei S, Morone F, Liljeros F, Makse H, Shaman JL. Inference and control of the nosocomial transmission of methicillin-resistant Staphylococcus aureus. eLife 7:e40977 (2018).

10. Pei S, SenPei-CU/COVID-19_US_Projection: COVID-19_US_Projection (https://github.com/SenPei-CU/COVID-19_US_Projection), Version 1, Zenodo (2020); http://doi.org/10.5281/zenodo.3722660.

